# Environmental impact and life cycle financial cost of hybrid (reusable/ single-use) instruments versus single-use equivalents in laparoscopic cholecystectomy

**DOI:** 10.1101/2021.03.10.21253268

**Authors:** Chantelle Rizan, Mahmood F Bhutta

**Author notes:** Corresponding author Address: ENT Department, Royal Sussex County Hospital, Eastern Road, Brighton, BN2 5BE. **Funding:** This work was funded by Surgical Innovations Ltd who manufacture hybrid laparoscopic instruments. However, the company played no part in scientific conduct, analysis, or writing of this manuscript.

## Abstract

**Background:** Hybrid surgical instruments contain both single-use and reusable components, potentially bringing together advantages from both approaches. The environmental and financial costs of such instruments have not previously been evaluated.

**Methods:** We used Life Cycle Assessment to evaluate the environmental impact of hybrid laparoscopic clip appliers, scissors and ports used for a laparoscopic cholecystectomy, comparing these with single-use equivalents. We modelled this using SimaPro and ReCiPe midpoint and endpoint methods to determine 18 midpoint environmental impacts including the carbon footprint, and three aggregated endpoint impacts. We also conducted life cycle cost analysis of products, taking into account unit cost, decontamination, and disposal costs.

**Results:** The environmental impact of using hybrid instruments for a laparoscopic cholecystectomy was lower than single-use equivalents across 17 midpoint environmental impacts, with mean average reductions of 60%. The carbon footprint of using hybrid versions of all three instruments was around one-quarter of single-use equivalents (1,756 g versus 7,194 g CO_2_e per operation), and saved an estimated 1.13 e^-5^ DALYs (disability associated life years, 74% reduction), 2.37 e^-8^ species.year (loss of local species per year, 76% reduction), and US $ 0.6 in impact on resource depletion (78% reduction). Scenario modelling indicated that environmental performance of hybrid instruments was better even if there was low number of reuses of instruments, decontamination with separate packaging of certain instruments, decontamination using fossil-fuel rich energy sources, or changing carbon intensity of instrument transportation. Total financial cost of using a combination of hybrid laparoscopic instruments was less than half that of single-use equivalents (GBP £131 versus £282).

**Conclusion:** Adoption of hybrid laparoscopic instruments could play an important role in meeting carbon reduction targets for surgery, and also save money.

## INTRODUCTION

The advent of minimally invasive surgery has led to huge advances in abdominal surgery over the last three decades, with advantages over open approaches including faster recovery, shortened hospital stay, and reduced pain and scarring.(1) An estimated 14 million laparoscopic procedures were performed worldwide in 2020, at which point the global laparoscopic devices and accessories market was estimated at US $ 13.7 billion per annum.(2) The most common laparoscopic procedure performed is cholecystectomy,(3) but others include appendicectomy, colectomy, and bariatric operations, as well as gynaecological and urological procedures.

Anecdotally many surgeons prefer single-use over reusable laparoscopic instruments due to historical concerns over sterility, or possible failure of reusable instruments (e.g. less reliably sharp dissecting scissors or failure of clip appliers).(4) However, the financial cost of single-use laparoscopic instruments is typically higher than reusables,(4) estimated at nineteen times more in laparoscopic cholecystectomy after accounting for costs of decontamination, repair, and replacement.(5) An additional factor in deciding which type of laparoscopic instrument to use should be the environmental impact, which has previously received little attention, but is an urgent agenda because of risks to planetary health. Healthcare is responsible for over 4% of global net greenhouse gas emissions,(6) and in England medical equipment is estimated to account for 10% of this,(7) with consumable items identified as a key carbon hotspot within the operating theatre.(8)

The most common metric for measuring environmental harm is the carbon footprint: an estimate of greenhouse gas emissions associated with a product, process or system, equated and summated as carbon dioxide equivalents (CO_2_e). A number of studies document that (almost universally) the carbon footprint of reusable items in the operating theatre is lower than that for single-use equivalents, including for scissors,(9) gowns and drapes,(10) laparotomy pads,(11) sharps containers,(12) and anaesthetic items (anaesthetic drug trays,(13) laryngeal mask airways,(14) and laryngoscope handles and blades).(15)

Hybrid instruments, also referred to as ‘resposable instruments’ or ‘modular systems’, are predominantly reusable, but with some single-use components. For example, in laparoscopy they may include a reusable trocar and port with disposable seal, or a reusable instrument handle with disposable insert. Such devices likely reduce the environmental impact as well as financial cost of laparoscopic instruments, but this has not previously been evaluated. Here we set out to compare the environmental and financial life-cycle cost of currently available hybrid instruments for laparoscopic cholecystectomy, and compared these to single-use equivalents.

## MATERIAL AND METHODS

### Functional unit

We included in our analysis three types of instrument routinely used in laparoscopic cholecystectomy (Table 1): laparoscopic clip appliers, laparoscopic scissors, and ports (small diameter 5mm ports, and large diameter 10-11mm ports). These instruments have both disposable and hybrid versions available on the market. The ‘functional unit’ (unit of analysis) was defined as the number of these three types of instruments typically required to perform one laparoscopic cholecystectomy; two small diameter ports, two large diameter ports, one laparoscopic scissor and one laparoscopic clip applier.

**Table 1:**
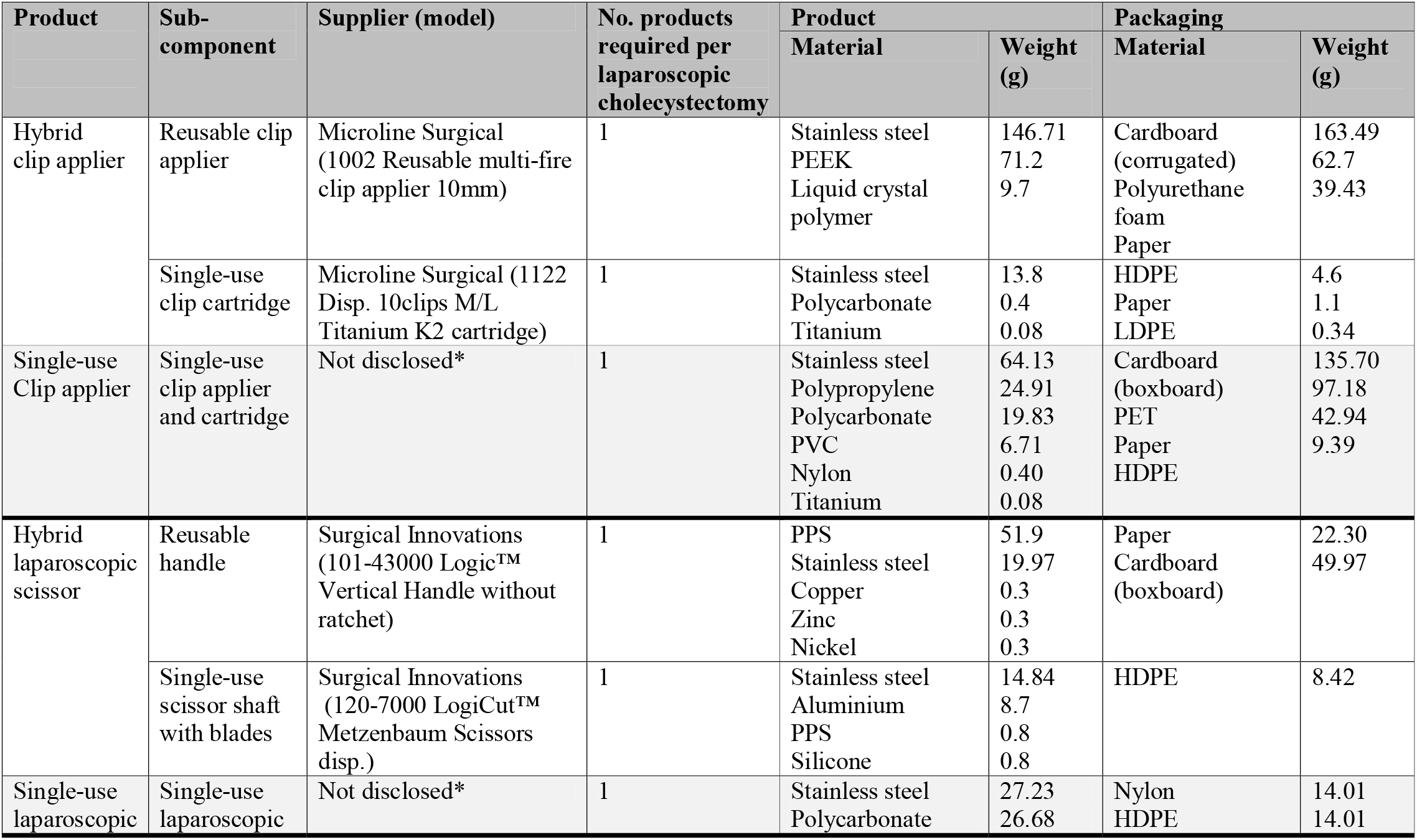
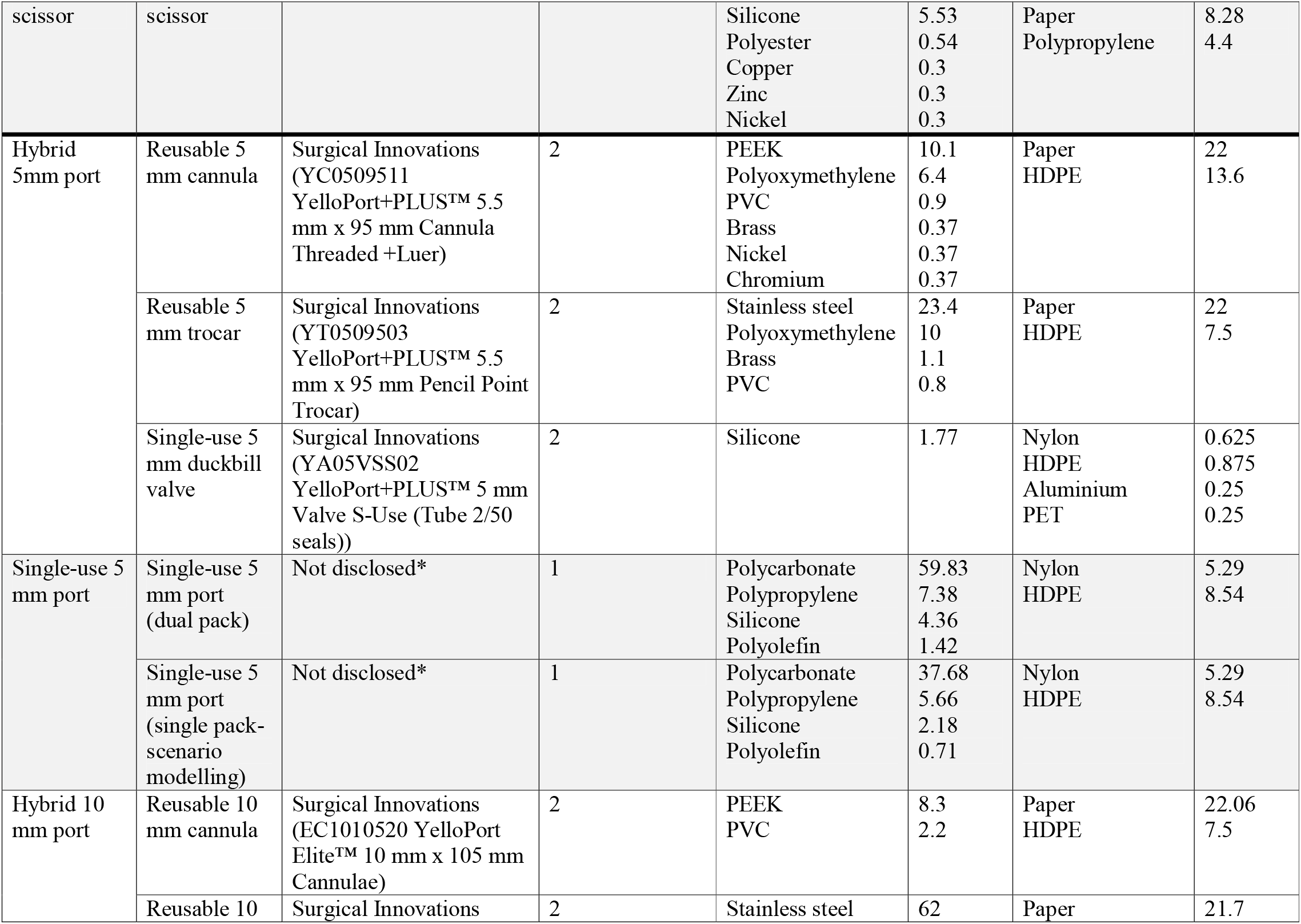
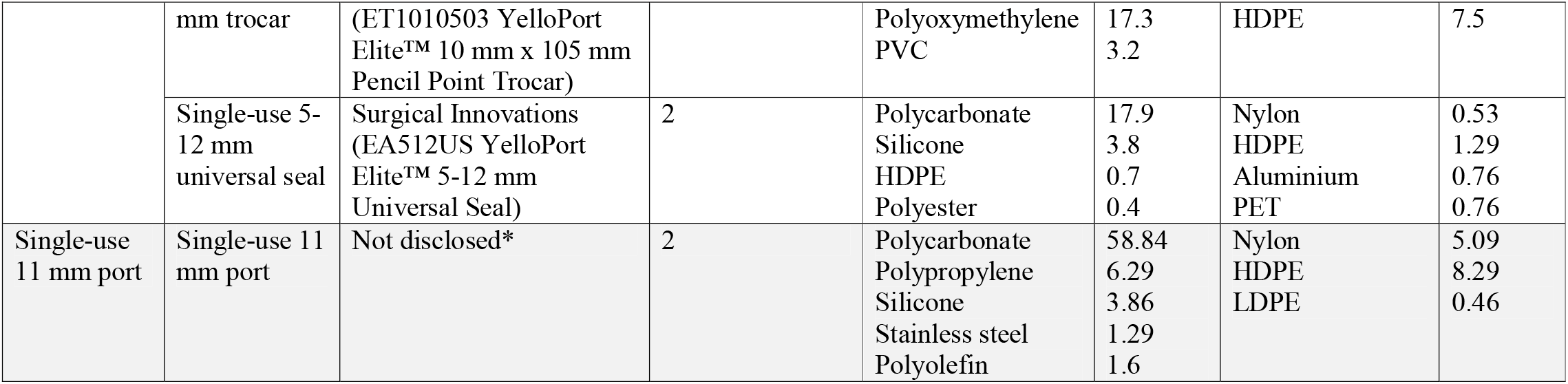
Material composition of laparoscopic clip applier, scissors, and ports. No.= number of uses (number of uses same for product and packaging), HDPE= high density polyethylene, LDPE= low density polyethylene; PEEK= polyether ether ketone, PET= Polyethylene terephthalate, PPS (polyphenylene sulphide), PVC= polyvinylchloride, * = not disclosed at request of manufacturers supplying information on material composition

### Determination of material composition of instruments for analysis

Hybrid instruments were supplied by Surgical Innovations Ltd (Leeds, UK) and Microline Surgical Inc (Beverly, USA). Equivalent single-use instruments were identified from the catalogue of the UK National Health Service (NHS) Supply Chain. We determined raw material composition of each instrument and associated primary packaging using information provided by manufacturers through personal correspondence, or expert knowledge where manufacturers were unable to provide sufficient detail. Weight of component materials was determined using Fisherbrand FPRS4202 Precision balance scales (Fisher Scientific, Loughborough, UK).

### Parameters for Life Cycle Assessment

An LCA was modelled using SimaPro v9.10 (PRé Sustainability, Amersfort, Netherlands), drawing upon ISO 14044 Guidelines.(16) We performed a ‘cradle to grave’ analysis, including raw material extraction, manufacture, transport, and disposal, plus decontamination for reusable components of hybrid instruments (system boundary outlined in Supplementary Figure 1). Other reusable instruments and consumables used to perform a laparoscopic cholecystectomy were beyond the scope of this analysis.

**Figure 1.**
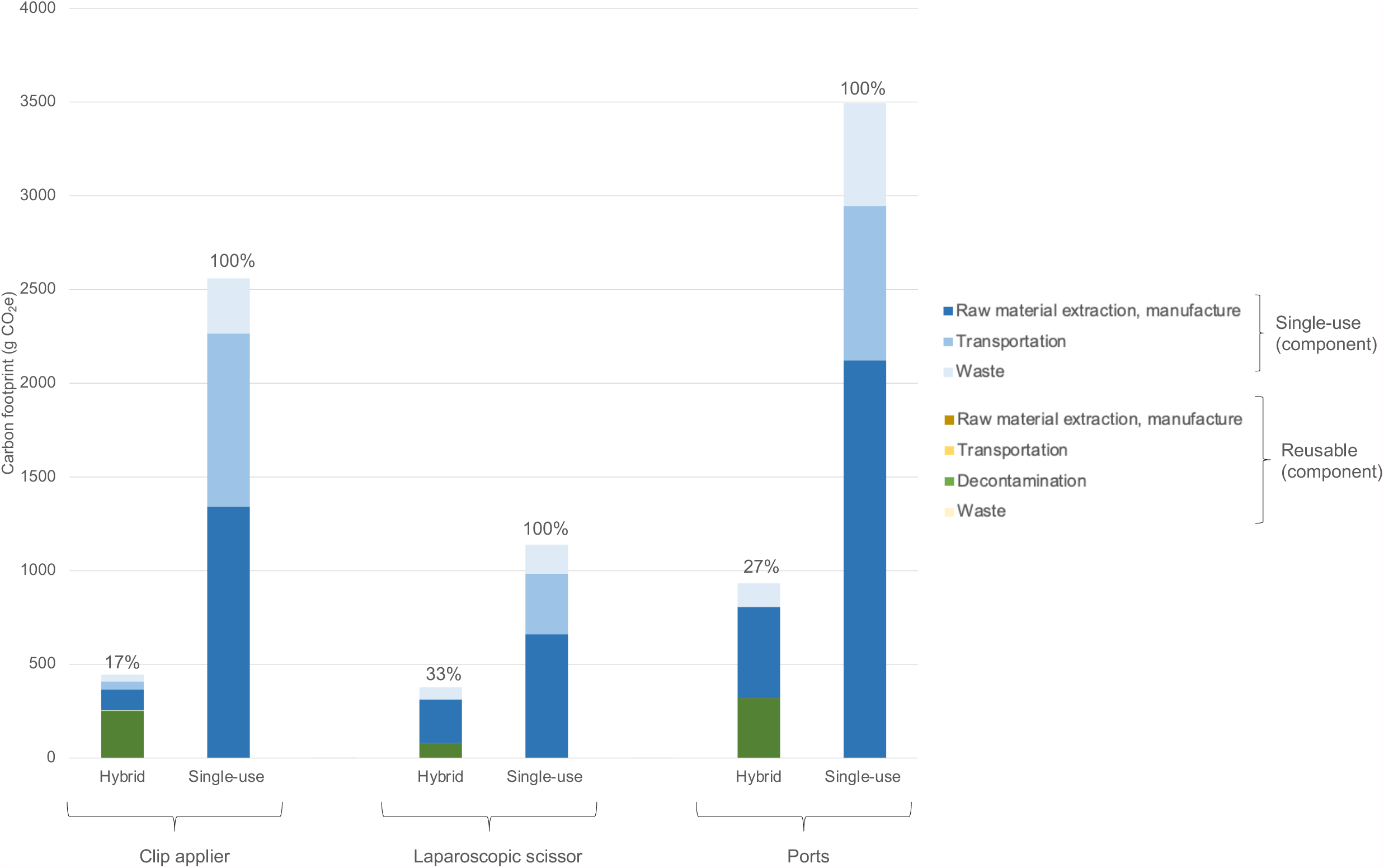
Carbon footprint of hybrid vs single-use laparoscopic clip applier, scissors, and ports, and relative contributions of sub-processes. Modelled per use of laparoscopic clip applier, one laparoscopic scissor, two 5mm ports, and two 10-11mm ports, comparing hybrid with single-use equivalents. Percentage figures above bars indicate proportion (%) relative to single-use equivalents. ‘Raw material extraction, manufacture’ includes global averages for transportation from site of extraction to the manufacturer.

Material specific global average environmental impact of raw material extraction, production, and transportation to the ‘end user’ (in this case the manufacturer) of instruments and packaging was determined through matching materials identified with the closest processes within Ecoinvent v3.6, or where unavailable in Industry data v.2.0 (both databases integrated within SimaPro).

Environmental impact of instrument manufacture was approximated using global average metal working processes for all metal components of instruments, and injection moulding processes for plastic components of instruments (as modelled in Ecoinvent v3.6). The mode and distance of international transportation from site of manufacture to the UK was determined through discussion with instrument suppliers (Supplementary Table 1), and we also assumed 80 km of travel by road using a heavy goods vehicle both within country of origin and in the UK, with the first and last 8 km at either end of this journey by courier.

All reusable components were assumed to be decontaminated and re-used 500 times, in accordance with manufacturer guidance on typical usage, with energy and material inputs for decontamination modelled using our own data presented elsewhere.(17) The metrics of the washer/ disinfector and sterilisation cycle will vary according to a number of factors, including the loading of machines, electricity source, and constitution of instrument sets. We assumed reusable components of hybrid instruments were integrated into a general laparoscopic set, as is common practice (Supplementary Table 2a). The typical weight of instruments on a general laparoscopic set was 730 g (data from our local hospital) and the weight of hybrid reusable components was 594g. Thus, hybrid instruments within a set comprise 45% of total weight (594/1324g), and so we also apportioned 45% of environmental harm from decontamination of the set to the reusable components of hybrid instruments. At the end of their life, all items were assumed to be disposed of as clinical waste via high-temperature incineration.

**Table 2:** Environmental impact (midpoint categories) per use of hybrid versus single-use laparoscopic clip applier, scissors, and ports. Modelled on use of one clip applier, one laparoscopic scissor, two 5mm ports, and two 10-11mm ports, comparing hybrid with single-use equivalents. Normalised results= environmental impact relative to the global average person’s contribution to the impact category over one year. 1,4-DCB =dichlorobenzene, CFC11= Trichlorofluoromethane, CO_2_e= carbon dioxide equivalents, Cu= copper, eq= equivalents, Bq Co-60 eq = becquerel Cobalt-60, m^2^a = square meter years, N= nitrogen, NO_x_= nitrous oxides, P=phosphate, PM2.5 = particulate matter <2.5 micrometres, SO_2_= sulphur dioxide

| Impact category | Unit | Laparoscopic clip applier |  | Laparoscopic scissors |  | Ports |  | Total (Normalised results) |  |
| --- | --- | --- | --- | --- | --- | --- | --- | --- | --- |
|  |  | Hybrid | Single-use | Hybrid | Single-use | Hybrid | Single-use | Hybrid | Single-use |
| Global warming | g CO<sub>2</sub>e | 445 | 2,559 | 378 | 1,139 | 933 | 3,495 | 1,756<br>(2.20e<sup>-4</sup>) | 7,194<br>(9.01e<sup>-4</sup>) |
| Stratospheric ozone depletion | g CFC11 eq | 0.0002 | 0.0008 | 0.0001 | 0.0005 | 0.0004 | 0.0013 | 0.0007<br>(1.16e<sup>-5</sup>) | 0.0026<br>(4.34e<sup>-5</sup>) |
| Ionizing radiation | Bq Co-60 eq | 57 | 80 | 28 | 25 | 79 | 59 | 164<br>(3.42e<sup>-4</sup>) | 164<br>(3.41e<sup>-4</sup>) |
| Ozone formation, Human health | g NO<sub>x</sub> eq | 0.89 | 8.12 | 0.79 | 3.17 | 1.42 | 8.21 | 3.10<br>(1.51e<sup>-4</sup>) | 19.51<br>(9.48e<sup>-4</sup>) |
| Fine particulate matter formation | g PM2.5 eq | 0.62 | 4.08 | 0.78 | 1.91 | 0.89 | 3.53 | 2.28<br>(8.93e<sup>-5</sup>) | 9.52<br>(3.72e<sup>-4</sup>) |
| Ozone formation, Terrestrial ecosystems | g NO<sub>x</sub> eq | 0.91 | 8.27 | 0.81 | 3.24 | 1.46 | 8.39 | 3.18<br>(1.79e<sup>-4</sup>) | 19.90<br>(1.12e<sup>-3</sup>) |
| Terrestrial acidification | g SO<sub>2</sub> eq | 1.18 | 8.53 | 1.44 | 4.46 | 2.08 | 8.91 | 4.70<br>(1.15e<sup>-4</sup>) | 21.90<br>(5.34e<sup>-4</sup>) |
| Freshwater eutrophication | g P eq | 0.12 | 0.62 | 0.17 | 0.26 | 0.17 | 0.43 | 0.46<br>(7.09e<sup>-4</sup>) | 1.31<br>(2.01e<sup>-3</sup>) |
| Marine eutrophication | g N eq | 0.09 | 0.06 | 0.04 | 0.05 | 0.12 | 0.07 | 0.24<br>(5.25e<sup>-5</sup>) | 0.18<br>(3.97e<sup>-5</sup>) |
| Terrestrial ecotoxicity | g 1,4-DCB | 3,976 | 19,767 | 5,628 | 8,939 | 1,171 | 4,142 | 10,776<br>(1.04e<sup>-2</sup>) | 32,849<br>(3.17e<sup>-2</sup>) |
| Freshwater ecotoxicity | g 1,4-DCB | 36 | 176 | 97 | 91 | 17 | 39 | 150<br>(1.23e<sup>-1</sup>) | 306<br>(2.5e<sup>-1</sup>) |
| Marine ecotoxicity | g 1,4-DCB | 47 | 230 | 122 | 118 | 23 | 54 | 192<br>(1.86e<sup>-1</sup>) | 402<br>(3.89e<sup>-1</sup>) |
| Human carcinogenic toxicity | g 1,4-DCB | 45 | 203 | 65 | 91 | 43 | 117 | 153<br>(5.52e<sup>-2</sup>) | 411<br>(1.48e<sup>-1</sup>) |
| Human non-carcinogenic toxicity | g 1,4-DCB | 576 | 2,871 | 952 | 1,386 | 390 | 1,013 | 1,919<br>(1.29e<sup>-2</sup>) | 5,269<br>(3.54e<sup>-2</sup>) |
| Land use | m<sup>2</sup>a crop eq | 0.01 | 0.16 | 0.01 | 0.02 | 0.02 | 0.03 | 0.04<br>(8.04e<sup>-6</sup>) | 0.21<br>(3.37e<sup>-5</sup>) |
| Mineral resource scarcity | g Cu eq | 9 | 39 | 14 | 19 | 3 | 4 | 26<br>(2.18e<sup>-7</sup>) | 62<br>(5.13e<sup>-7</sup>) |
| Fossil resource scarcity | g oil eq | 137 | 784 | 100 | 315 | 261 | 940 | 498<br>(5.08e<sup>-4</sup>) | 2,039<br>(2.08e<sup>-3</sup>) |
| Water consumption | m<sup>3</sup> | 0.0030 | 0.0146 | 0.0028 | 0.0083 | 0.0081 | 0.0208 | 0.0139<br>(5.22e<sup>-5</sup>) | 0.0437<br>(1.64e<sup>-4</sup>) |

Processes selected from SimaPro databases are detailed in Supplementary Table 3.

**Table 3.** Environmental impact (endpoint categories) of hybrid versus single-use laparoscopic clip applier, scissors, and ports. Environmental impacts (midpoint categories) measured using life cycle assessment and modelled on one laparoscopic clip applier, one laparoscopic scissor, two 5mm ports, and two 10-11mm ports (one use; number required to perform a single laparoscopic cholecystectomy), comparing hybrid with single-use equivalents. Normalised results= environmental impact relative to the global average person’s contribution to the impact category over one year. DALYs= disability adjusted life years, species.year=loss of local species per year, US$ =extra costs involved for future mineral and fossil resource extraction

| Damage category | Unit | Laparoscopic clip applicator |  | Laparoscopic scissors |  | Ports |  | Total (Normalised results) |  |
| --- | --- | --- | --- | --- | --- | --- | --- | --- | --- |
|  |  | Hybrid | Single-use | Hybrid | Single-use | Hybrid | Single-use | Hybrid | Single-use |
| Human health | DALY | 1.09e<sup>-6</sup> | 6.30e<sup>-6</sup> | 1.28e<sup>-6</sup> | 2.90e<sup>-6</sup> | 1.67e<sup>-6</sup> | 6.13e<sup>-6</sup> | 4.04e<sup>-6</sup><br>(1.7e<sup>-4</sup>) | 1.53e<sup>-5</sup><br>(6.45e<sup>-4</sup>) |
| Ecosystems | species.yr | 1.96e<sup>-9</sup> | 1.24e<sup>-8</sup> | 1.84e<sup>-9</sup> | 5.22e<sup>-9</sup> | 3.67e<sup>-9</sup> | 1.36e<sup>-8</sup> | 7.47e<sup>-9</sup><br>(1.04e<sup>-5</sup>) | 3.12e<sup>-8</sup><br>(4.36e<sup>-5</sup>) |
| Resources | US \$ | 0.0464 | 0.2944 | 0.0314 | 0.1176 | 0.0853 | 0.344473 | 1.63e<sup>-1</sup><br>(5.82e<sup>-6</sup>) | 7.56e<sup>-1</sup><br>(2.7e<sup>-5</sup>) |

### Assessment of environmental impact

Following development of the LCA inventory within SimaPro (which identifies flow of energy, materials and water to and from nature resulting from relevant processes), we used the ReCiPe v1.1 Midpoint Hierarchist method (integrated within SimaPro) to characterise such emissions, and to combine these into environmental impacts. This method evaluates eighteen midpoint impact categories (each relating to a single environmental problem): global warming, stratospheric ozone depletion, ionising radiation, ozone formation (on human health, and terrestrial ecosystems), fine particulate matter formation, terrestrial acidification, eutrophication (freshwater and marine), ecotoxicity (terrestrial, freshwater, and marine), human toxicity (carcinogenic and non-carcinogenic), land use, resource scarcity (mineral and fossil), and water consumption. We performed deeper analysis of global warming impact, and conducted hotspot analysis to determine processes contributing most to carbon footprints. We used the ReCiPe v1.1 Endpoint Hierarchist method to aggregate midpoint impact categories to calculate endpoint factors for damage to human health, the natural environment, and resource scarcity. Finally, we used ReCiPe v1.1 Hierarchist normalisation factors to compare total midpoint and endpoint impacts to mean average contributions to each of those impacts from a global average person’s daily routine activities.(18)

### Scenario modelling

To determine sensitivity of results to allocation methods and key assumptions, we modelled five alternative scenarios.

First, we determined the impact of altering the number of uses of instruments, and through this also identified the threshold at which the carbon footprint of reusables became lower than using single-use equivalents.

Laparoscopic clip appliers are not used across all laparoscopic operations performed in a given surgical department (but laparoscopic scissors and ports typically are), and clip appliers are therefore not always integrated into general laparoscopic sets. In our second scenario we modelled decontamination with the clip applier decontaminated separately in a flexible double wrapped polyethylene pouch (Supplementary Table 2b). Here we assumed total weight of items in the laparoscopic general set was 1.1kg, with decontamination of surgical scissors and ports apportioned accordingly (366/1096 g = 33%).

Third, we modelled the impact of switching the electricity source for decontamination to that typical of Australia, a country using a lower proportion of renewable energy.

Fourth, we modelled impact of changing overseas transport of single-use instruments to shipping by sea, with distances determined using the online Pier2Pier tool (19) and alternative road distances using Google maps(20) (Supplementary Table 1).

Finally, we modelled the carbon footprint of using three 5mm ports and one 10/11mm port, as this is a commonly used alternative port configuration for laparoscopic cholecystectomy. In the baseline scenario we modelled 5mm single-use ports based upon a dual pack (containing two cannulas, one syringe, and one trocar), and for this alternative scenario, a single-pack was modelled based on removal of duplicate components (Table 1).

### Consequentialist approach to LCA

Our baseline analysis uses standard practice of an attributional approach to LCA,(21) where products are allocated a proportion of the total lifecycle material and energies required for processes shared with other products. We performed alternative analysis of our base scenario using the consequential approach to LCA, which examines consequences of a change.(21) Specifically, when switching from single-use to hybrid laparoscopic instruments, hybrid components are integrated into surgical sets already destined for decontamination, meaning under the consequentialist model there is no additional impact from decontamination.

### Assessment of life cycle financial cost

The cost of instruments reported in the NHS Supply Chain database(22) was equated to cost of manufacture and distribution. Disposal at the end of instrument life was costed at £ 617.22 per tonne, based on the price of clinical waste incineration reported by the NHS Digital Estates Returns Information Collection dataset.(23) Cost of decontaminating reusable components of hybrid instruments was based on the charge made by our hospital sterilisation services per instrument set, apportioned according to the weight of hybrid reusable components (here 45%).

### Role of the funding source and ethical considerations

This work was funded by Surgical Innovations Ltd who manufacture hybrid laparoscopic instruments, but the company played no part in scientific conduct, analysis, or writing of this manuscript. This study did not require ethical approval or consent as there were no patients or participants involved in this research.

## RESULTS

### Environmental life cycle assessment

Table 1 details material composition and life-span assumptions used for the laparoscopic instruments. The carbon footprint per operation of a laparoscopic hybrid instrument compared to its single-use equivalent was 17% for a clip applier (445 g versus 2,559 g CO_2_e); 33% for scissors (378 g versus 1,139 g CO_2_e); and 27% for four ports (933 g CO_2_e versus 3,495 g CO_2_e/ operation) (Figure 1). When combined, the carbon footprint of using hybrid versions of all three instrument types for an operation was 24% of that of single-use equivalents (1,756 g CO_2_e versus 7,194 g CO_2_e), saving a total of 5.4 kg CO_2_e. This equates to the normal activities of a global average person over 6 hours (normalised results).

Hotspot analysis indicated that the majority of the carbon footprint of hybrid instruments was due to the single-use components (mean 62%, range 43% -79%), followed by decontamination of reusable components (mean: 37%, range 21-56%) (Supplementary Table 4). For single-use instruments, raw material extraction and manufacture (including transportation between these two processes) was a major contributor (mean 57%, range 52% -61%), followed by onward transportation (mean 29%, range 24%-36%), and waste (mean 14%, range 12% -16%).

**Table 4.**
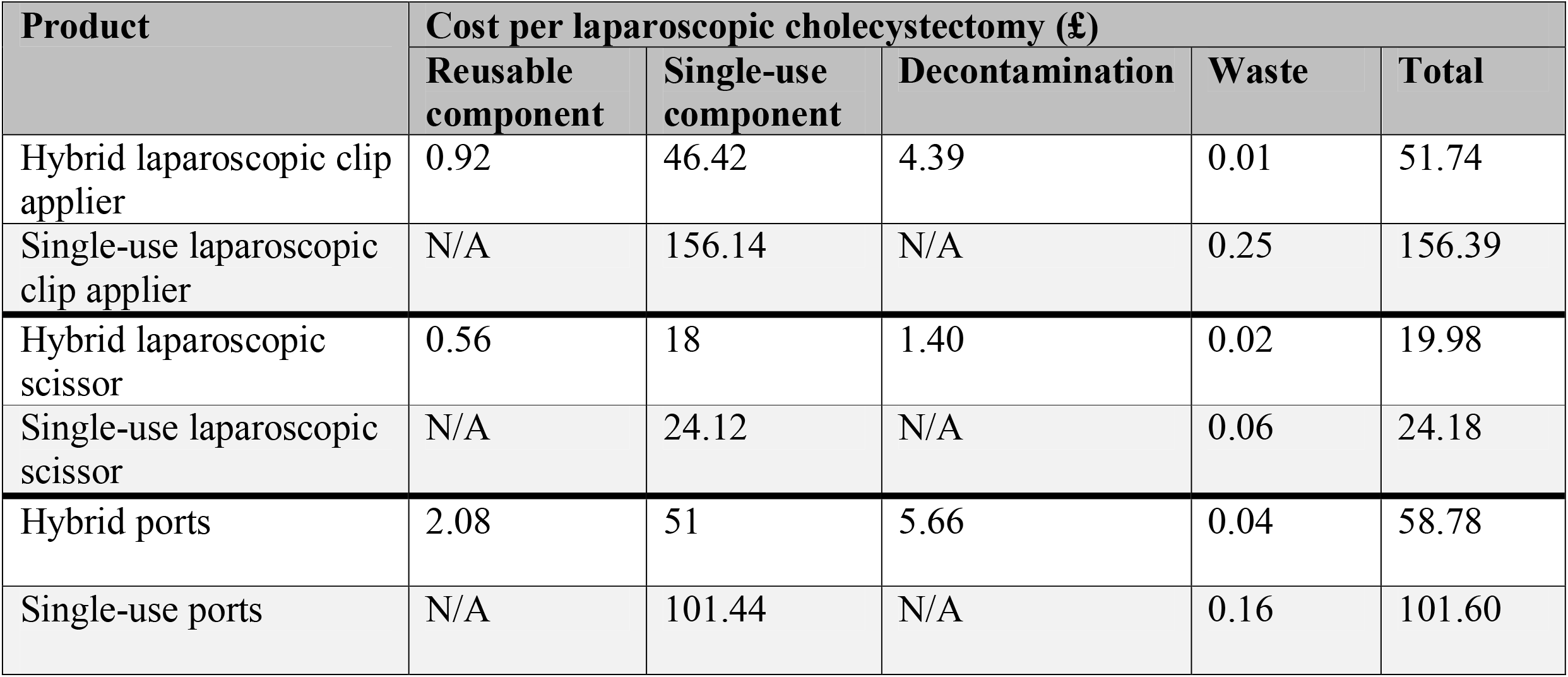
Life cycle costing of hybrid vs single-use laparoscopic clip applier, scissors, and ports. Cost per laparoscopic cholecystectomy, modelled on one clip applier, one laparoscopic scissor, two 5mm ports, and two 10-11mm ports, comparing hybrid with single-use equivalents

The environmental impact of using a combination of hybrid laparoscopic clip appliers, scissors, and ports for an operation was lower than using single-use equivalents across all 18 midpoint environmental impacts except for marine eutrophication (Supplementary Figure 2), with mean average reductions in environmental impact of 60% (range -32% to 84%). Disaggregated data for each instrument showed midpoint environmental impact categories to be lower for each hybrid product, with a small number of exceptions (Table 2). Contribution analysis (Supplementary Figures 3 and 4) indicated that the 3% higher freshwater ecotoxicity and 7% higher marine ecotoxicity impacts for hybrid compared to single-use laparoscopic scissors were principally due to aluminium within the single-use component of the hybrid scissors (38% for freshwater ecotoxicity, and 37% for marine ecotoxicity), and due to copper assumed to be used within the metal working process (contributing 50% to both categories). Higher marine eutrophication impacts for the hybrid laparoscopic clip applier (38% higher) and ports (76% higher) were largely attributable (89% and 85% respectively) to the handling of wastewater from decontamination (Supplementary Figure 5 and 6). The ionising radiation impact of hybrid laparoscopic scissors and ports (Supplementary Figure 7 and 8) was higher than single-use equivalents (14% and 32% respectively) due to the electricity used in the decontamination process (accounting for 55% and 78% of the impact respectively).

For endpoint categories, using a combination of hybrid laparoscopic clip appliers, scissors, and ports for a single laparoscopic cholecystectomy saved an estimated 1.13 e^-5^ DALYs (disability associated life years), 2.37 e^-8^ species.year (loss of local species per year), and US $ 0.6 impact on resource depletion, representing reductions of 74%, 76% and 78% respectively compared to single-use equivalents (Table 3, Supplementary Figure 9).

### Scenario modelling

For all hybrid instruments, carbon footprint was lower than single-use equivalents when the reusable component was used more than twice. Impact on carbon plateaued at around 10 uses of reusable components, with little additional gain (<1%) after using laparoscopic scissors 60 times, ports 70 times, and clip appliers 100 times (Supplementary Figure 10). However, continued use of these saves the additional carbon burden of obtaining new instruments.

When packaged and decontaminated separately, the carbon footprint of the hybrid laparoscopic clip applier increased 3.7 fold to 1,650 g CO_2_e per use (Supplementary Table 5). There were small accompanying increases for laparoscopic scissors (to 394 g CO_2_e per use, 4% increase) and ports (999 g CO_2_e per use, 7% increase), due to a greater proportional weight in the instrument set. Nevertheless, in this alternative model the carbon footprint of all hybrid instruments remained lower than single-use equivalents (36% less for laparoscopic clip appliers, 65% less for laparoscopic scissors, and 71% less for ports).

The carbon footprint of the decontamination process itself was 54% higher when Australian electricity was modelled, which increased carbon footprint of the hybrid instruments by 11% - 30% (Supplementary Table 6), but this remained lower (63%-77%) than single-use equivalents.

Shipping in place of air freight for international transport of single-use instruments reduced carbon footprint by 22% -33% relative to the baseline single-use items (Supplementary Table 7), but the hybrid baseline instruments remained lower than the shipped single-use equivalents; by 74% for the clip applier, 55% for scissors, and 65% for ports.

Finally, using three hybrid 5mm ports and one 10mm port (635 g CO_2_e/ operation) resulted in a 32% reduction in carbon footprint relative to the base scenario hybrid port setup (Supplementary Table 8). The use of single-use ports with this alternative port configuration was associated with six-fold increase in carbon footprint compared with use of hybrids (3,613 g CO_2_e), constituting a 3% increase relative to the base scenario single-use port setup.

### Consequentialist approach to LCA

Under the consequential approach to LCA, carbon footprint of a hybrid laparoscopic clip applier was 198 g CO_2_e (7% of single-use equivalent of 2,559 g CO_2_e), for scissors this was 299 g CO_2_e (26% of single-use equivalent of 1,139 g CO_2_e), and for four hybrid ports was 614 g CO_2_e (18% of single-use equivalent of 3,495 g CO_2_e). When combined, under the consequentialist approach the carbon footprint of using hybrid versions of all three instruments types for an operation was 15% of that for single use equivalents (1,110 g versus 7,194 g CO_2_e), saving a total of 6,083 g CO_2_e.

### Life cycle financial cost

Per operation, the cost of hybrid compared to single-use instruments (Table 4) was 33% for a clip applier (GBP £ 52 versus £ 156), 83% for scissors (GBP £ 20 versus £ 24), and 58% for ports (GBP £ 59 versus £ 102). Most of these costs (≥87% contribution in all cases) were from single-use items or components, with smaller contributions from decontamination (≤10%), reusable components (≤ 4%), or waste (≤ 0.2%). For a single laparoscopic cholecystectomy, cost of using a combination of hybrid laparoscopic clip appliers, scissors and ports was 47% of that of single-use equivalents (GBP £ 131 versus £ 282).

## DISCUSSION

The NHS has recently set the ambitious aim to become the world’s first ‘net zero’ carbon national health service.(7) Reducing the number of single-use items used to deliver surgical care, and opting for low carbon alternatives within the operating theatre will be important in meeting that ambition.

Here we found that the carbon footprint of using hybrid scissors, ports and clip appliers was 76% lower than using single-use equivalents, saving 5.4 kg CO_2_e per operation (equal to driving 16 miles in an average petrol car). There are almost 73,000 laparoscopic cholecystectomy operations performed each year in England,(24) and if the three hybrid instruments analysed were used across all of these operations in place of single-use equivalents, this would save 396 tonnes CO_2_e, equivalent to 50 years of daily activities of a global average person (normalised results), or driving 1.2 million miles in an average petrol car.(25) Additional annual environmental benefits would be an estimated 0.82 DALYs (equating to 296 disability adjusted life days), 0.0017 species.year, and US $ 43,188 due to resource depletion. Under a consequentialist analysis, these values would be even higher. The direct financial costs across the lifespan of the hybrid instruments were less than half those of single-use items, and would save over £ 11 million per year if adopted for all laparoscopic cholecystectomies in England.

Scenario analysis indicated that use of three 5mm hybrid ports and one 10mm port (instead of two of each), reduced carbon footprint by around one-third, principally due to the difference in weight of the single-use component (10 mm hybrid port was associated with a 22.8 g single-use universal seal, whilst the 5mm hybrid port was associated with a 1.77 g single-use valve). Adoption of this downsized hybrid port setup may therefore correspond with additional environmental benefits, with further reductions anticipated for three-port microlaparoscopic approaches, eliminating the need for one of the ports altogether. However, adoption of the alternative port setup was associated with marginal increases for single-use ports when obtained as three individually wrapped 5mm ports (each containing its own syringe, trocar, and cannula), and one 11mm port. This contrasts with use of a double pack containing two 5mm ports, as modelled in the base scenario, where one syringe and one trocar was shared between two cannulas. This indicates that where single-use ports are used, use of dual packs (and development of triple packs for surgeons wishing to use this configuration), may confer environmental benefit where this eliminates duplicate single-use items.

Use of plastics within healthcare is also gaining increasing attention,(26) and in separate analyses (data not shown) we found that when using hybrid laparoscopic clip appliers, scissors, and ports for a laparoscopic cholecystectomy, the total plastic was 15% that of using single-use equivalents, and generated around 15% of the waste. If translated across all laparoscopic cholecystectomies in England this would save an estimated production and disposal of 30 tonnes of plastic per year.

The carbon footprint of a laparoscopic cholecystectomy has not previously been estimated and is beyond the scope of this study, but the carbon footprint of the CO_2_ gas used for abdominal insufflation in laparoscopic or other procedures has been estimated at 141 kg CO_2_e per operation,(27) and the carbon footprint of a laparoscopic endometrial staging procedure at 29.2 kg CO_2_e,(28) and a laparoscopic hysterectomy at 562 kg CO_2_e.(29) The wide variation in these figures is likely due to differences in study methods, boundaries, and assumptions, but nevertheless indicate that switching to hybrid laparoscopic instruments in place of single-use equivalents could significantly impact the overall carbon footprint of a laparoscopic procedure.

The principle of using hybrid in preference to single-use instruments and of minimising single-use components is likely generalisable to other hybrid laparoscopic instruments and other laparoscopic procedures. We are aware of hybrid laparoscopic articulating dissectors, retractor rings, electrocautery probes, and a variety of forceps on the market, and further research and innovation may expand this repertoire to enable further replacement of single-use equivalents. However, we also recognise that entirely re-useable instruments will (and should) always remain preferable to hybrid equivalents.

As with all LCAs, our analysis is limited by assumptions, the system boundary (products and processes included), parameter uncertainty (potential inaccuracy in the primary activity data, and emission factors embedded within SimaPro), and model uncertainty (limitation of the extent to which our model reflects reality). Nevertheless, our scenario modelling found hybrid laparoscopic instruments remained preferable to single-use equivalents even if there was infrequent reuse of hybrid instruments, if hybrid clip appliers were packaged and decontaminated separately to the main instrument set, if using a fossil-fuel rich source for decontamination, if switching international transport of single-use equipment to shipping in preference to air-freight, or if altering port configuration. Nevertheless, to maximise environmental benefits of hybrid instruments they should be used for their full lifespan, decontaminated within main instrument sets where possible, and manufactured, transported, and decontaminated using low carbon intensity methods.

We acknowledge that there are alternative solutions to the products evaluated, beyond the scope of this study. Given that a mean average of two-thirds of the carbon footprint of hybrid instruments was due to the single-use components, it is likely that increasing the reusable proportion of products would improve environmental impact. However, we are aware of suboptimal anecdotal user experience associated with current hybrid solutions with higher reusable portions, for example delays due to reloading of reusable clip appliers with single polymer or titanium locking clips (contrasting with cartridge containing multiple clips modelled here), and higher levels of technical skills required to use a reusable pre-tied knot pusher. Whilst fully reusable laparoscopic scissors exist, these are reportedly less reliably sharp than single-use equivalents. Future innovation should therefore be targeted towards improving design of reusable laparoscopic equipment, minimising the single-use component as far as possible, and scheduling maintenance and repair for reusable components.

## Conclusion

The carbon footprint of using hybrid instruments for laparoscopic cholecystectomy is around a quarter of that for single-use equivalents, and the financial cost around half. Given the global scale of laparoscopic surgery, adoption of hybrid instruments could play an important role in meeting carbon reduction targets in healthcare, and saving money.

## Data Availability

Supporting data is available as supplementary information. For further information contact the corresponding author

## Acknowledgements

We thank Surgical Innovations Ltd who provided data on the hybrid instruments, and some data on single-use equivalents. We thank other industry partners who provided data, but wish to remain anonymous. We also thank Miss Victoria Pegna for her specialty specific input.

## References

1. Agha R, Muir G (2003) Does laparoscopic surgery spell the end of the open surgeon? J R Soc Med 96(11):544–6.

2. iData Research (2020) Laparoscopic Devices Market Analysis, Size, Trends. Available at: https://idataresearch.com/product/laparoscopic-devices-market/. Accessed 27 Jan 2021.

3. Comitalo JB (2012) Laparoscopic cholecystectomy and newer techniques of gallbladder removal. JSLS 16(3):406–12.

4. Siu J, Hill AG, MacCormick AD (2017) Systematic review of reusable versus disposable laparoscopic instruments: costs and safety. ANZ J Surg 87(1-2):28–33.

5. Adler S, Scherrer M, Rückauer KD, Daschner FD (2005) Comparison of economic and environmental impacts between disposable and reusable instruments used for laparoscopic cholecystectomy. Surg Endosc 19(2):268–72.

6. Health Care without Harm (2019) Health care’s climate footprint, Climate-smart health care series green paper number one. Available at: https://noharm-uscanada.org/content/global/health-care-climate-footprint-report. Accessed 27 Jan 2021.

7. NHS England and NHS Improvement (2020) Delivering a ‘Net Zero’ National Health Service. Available at: https://www.england.nhs.uk/greenernhs/wp-content/uploads/sites/51/2020/10/delivering-a-net-zero-national-health-service.pdf. Accessed 27 Jan 2020.

8. Rizan C, Steinbach I, Nicholson R, Lillywhite R, Reed M, Bhutta MF (2020) The carbon footprint of operating theatres: a systematic review. Annals of Surgery 272(6):986–95.

9. Ibbotson S, Dettmer T, Kara S, Herrmann C (2013). Eco-efficiency of disposable and reusable surgical instruments-a scissors case. The International Journal of Life Cycle Assessment 18:1137–48.

10. Overcash M (2012). A comparison of reusable and disposable perioperative textiles: sustainability state-of-the-art. Anesth Analg 114(5):1055–66.

11. Kummerer K, Dettenkofer M, Scherrer M (1996) Comparison of reusable and disposable laparotomy pads. The International Journal of Life Cycle Assessment 1(2):67–73.

12. McPherson B, Sharip M, Grimmond T (2019) The impact on life cycle carbon footprint of converting from disposable to reusable sharps containers in a large US hospital geographically distant from manufacturing and processing facilities. PeerJ 7:e6204.

13. McGain F, McAlister S, McGavin A, Story D (2010) The financial and environmental costs of reusable and single-use plastic anaesthetic drug trays. Anaesth Intensive Care 38(3):538–44.

14. Eckelman M, Mosher M, Gonzalez A, Sherman J (2012) Comparative life cycle assessment of disposable and reusable laryngeal mask airways. Anesth Analg 114(5):1067–72.

15. Sherman JD, Raibley LA, Eckelman MJ (2018) Life Cycle Assessment and Costing Methods for Device Procurement: Comparing Reusable and Single-Use Disposable Laryngoscopes. Anesth Analg 127(2):434–43.

16. International Organization for Standardazation (2016). ISO 14044: 2006 Environmental management-Life cycle assessment-Requirements and guidelines. Available at: https://www.iso.org/standard/38498.html. Accessed 27 Jan 2021.

17. Rizan C, Bhutta M, Reed M, Lillywhite R (2020). The carbon footprint of processing reusable surgical instruments. Life Cycle Innovation Conference; Berlin, Germany. Available at: https://fslci.org/groups/lcic-2020/forum/topic/discussion-session-13-sustainable-pathways-for-decarbonization/. Accessed 27 Jan 2021.

18. Sleeswijk AW, van Oers LF, Guinée JB, Struijs J, Huijbregts MA. Normalisation in product life cycle assessment: an LCA of the global and European economic systems in the year 2000. Sci Total Environ. 2008;390(1):227–40.

19. Pier2Pier. Pier2Pier. 2021. http://www.pier2pier.com/Co2/ (accessed Mar 5 2021).

20. Google. Google Maps. 2021. https://www.google.com/maps (assessed Mar 5 2021).

21. Institute of World Resources, 2011 Greenhouse gas protocol, product life cycle accounting and reporting standard. https://www.wri.org/publication/greenhouse-gas-protocol-product-life-cycle-accounting-and-reporting-standard. 2011. (accessed 27 Dec 2019).

22. NHS Supply Chain (2017) NHSSC Minimally Invasive FAG16308-01.02.17-31.01.21. [Database]. Accessed 27 Jan 2021.

23. NHS Digital (2020) Estates Return Information Collection (ERIC) for 2019/20. Available at: https://digital.nhs.uk/data-and-information/publications/statistical/estates-returns-information-collection/england-2019-20. Accessed 27 Jan 2021.

24. NHS Digital (2020) Hospital Admitted Patient Care Activity, 2019-20. Available at: https://digital.nhs.uk/data-and-information/publications/statistical/hospital-admitted-patient-care-activity/2019-20. Accessed 27 Jan 2021.

25. Department for Environment Food and Rural Affairs (2020) Greenhouse gas reporting: conversion factors 2020. Available at: https://www.gov.uk/government/publications/greenhouse-gas-reporting-conversion-factors-2020. Accessed 27 Jan 2021.

26. Rizan C, Mortimer F, Stancliffe R, Bhutta MF (2020) Plastics in healthcare: time for a re-evaluation. J R Soc Med 113(2):49–53.

27. Power NE, Silberstein JL, Ghoneim TP, Guillonneau B, Touijer KA (2012) Environmental impact of minimally invasive surgery in the United States: an estimate of the carbon dioxide footprint. J Endourol 26(12):1639–44.

28. Woods DL, McAndrew T, Nevadunsky N, Hou JY, Goldberg G, Yi-Shin Kuo D, et al (2015) Carbon footprint of robotically-assisted laparoscopy, laparoscopy and laparotomy: a comparison. Int J Med Robot 11(4):406–12.

29. Thiel CL, Woods NC, Bilec MM (2018) Strategies to Reduce Greenhouse Gas Emissions from Laparoscopic Surgery. Am J Public Health 108(S2):S158–S64.

