## Supplementary figure for "Environmental impact and life cycle financial cost of hybrid (reusable/ single-use) instruments versus single-use equivalents in laparoscopic cholecystectomy"

**Supplementary Figure 1: system boundary**

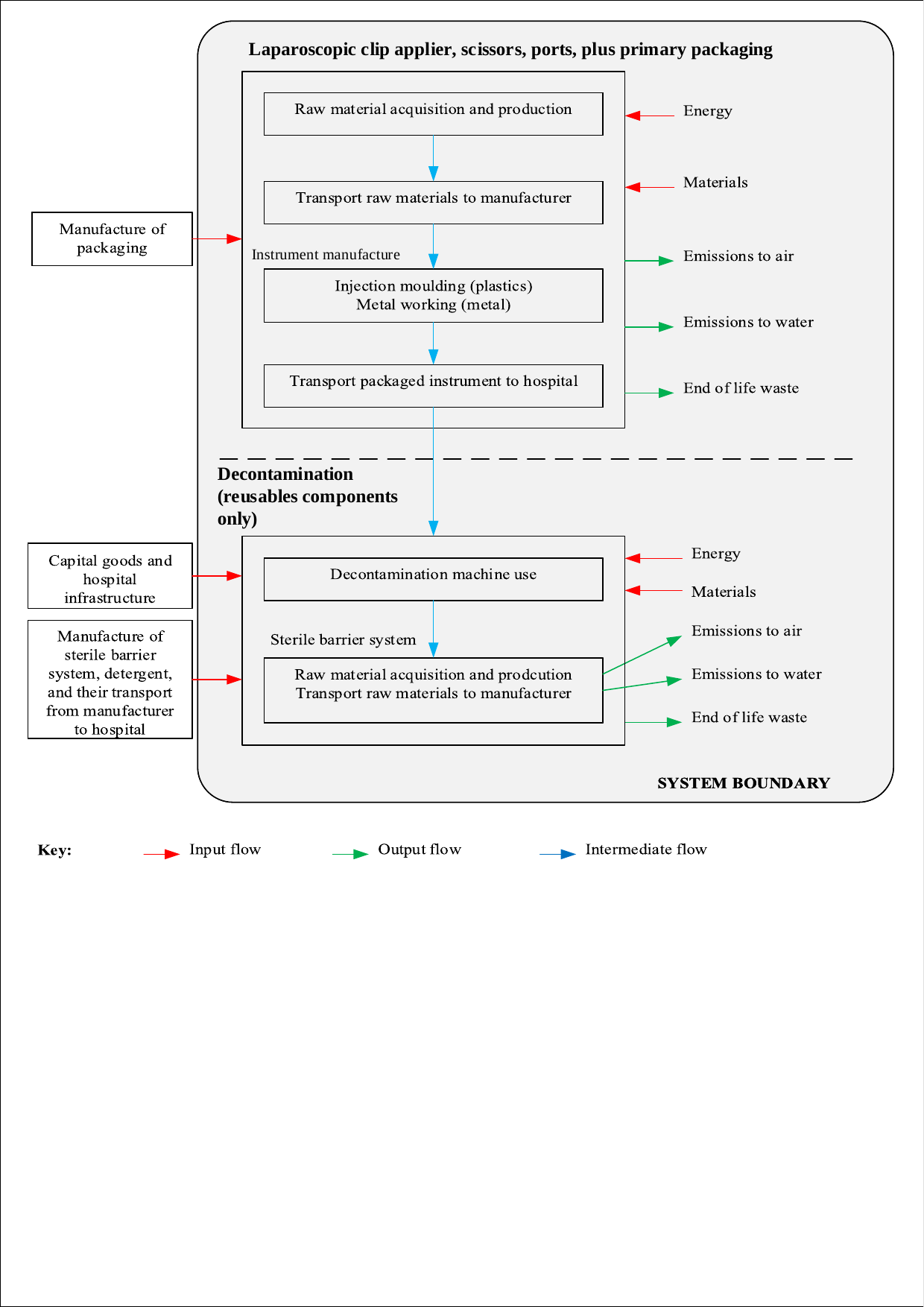

System boundary for life cycle assessment (LCA) of hybrid and single-use instruments (laparoscopic clip applier, scissors, and ports, and associated primary packaging). Processes included were energy and materials required for raw material acquisition, production and transport of instruments, alongside injection moulding (for plastic components) and metal working (for metal components). The manufacture of associated packaging was excluded. Processes included in decontamination (for the reusable components of hybrid instruments only), were energy and materials required by the decontamination machines, and for the sterile barrier system (including associated raw material acquisition and production, and transport of raw materials to the manufacturer). Capital goods and the hospital infrastructure were excluded, alongside manufacture and transport from the manufacturer to hospital of the sterile barrier system and detergent. Emissions to air and water were included for all processes excluding the decontamination machine. End of life waste was included for all materials.

**Supplementary Figure 2: Environmental impact (midpoint categories) of hybrid vs single-use laparoscopic clip applier, scissors, and ports**

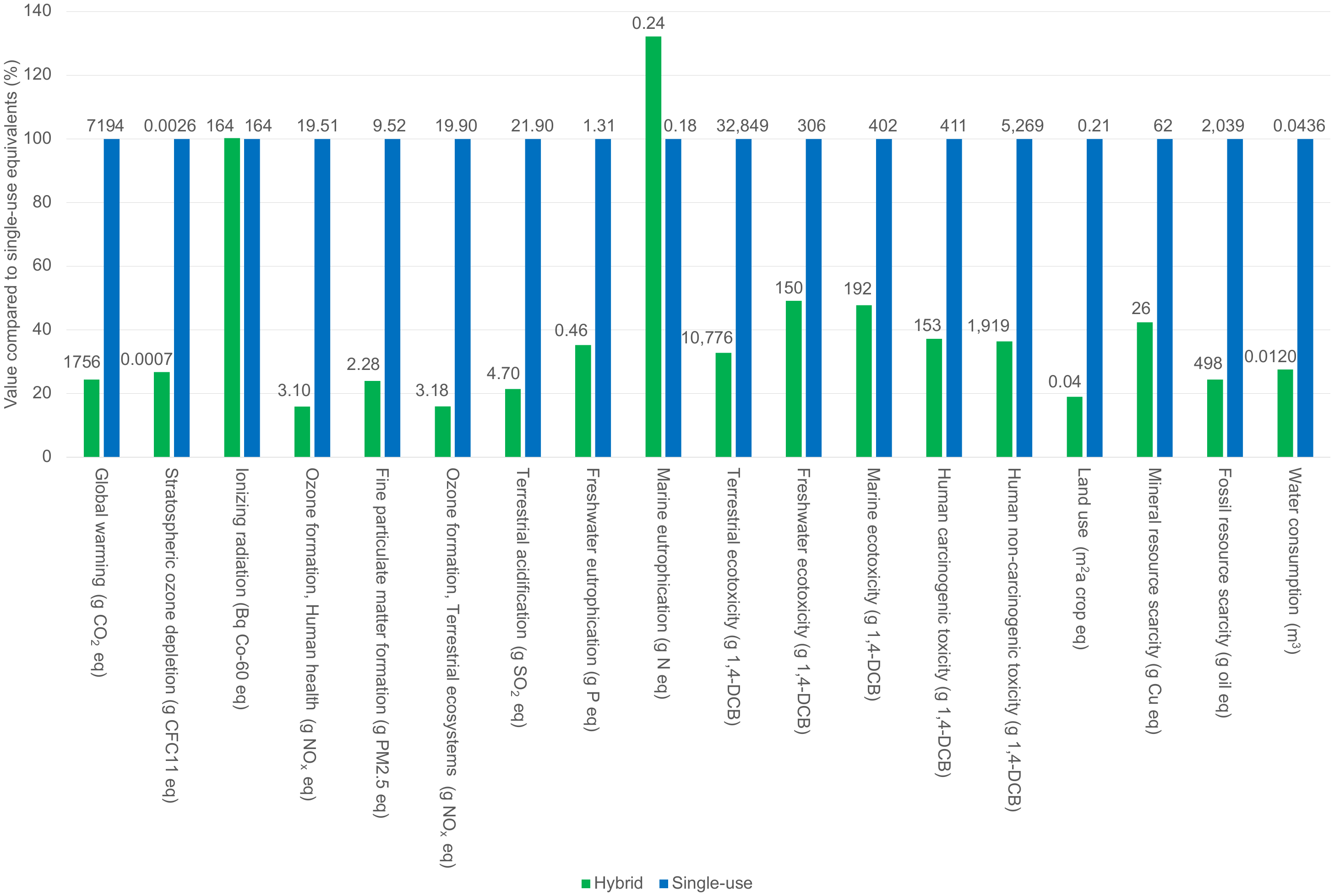

Modelled on one laparoscopic clip applier, one laparoscopic scissor, two 5mm ports, and two 10-11mm ports (one use; number required to perform a single laparoscopic cholecystectomy), comparing hybrid with single-use equivalents. Environmental impact compared as proportion (%) of single-use equivalents. Number above bars relate to midpoint category absolute figures for product. 1,4-DCB =dichlorobenzene, CFC11= Trichlorofluoromethane, CO_2_e= carbon dioxide equivalents, Cu= copper, eq= equivalents, Bq Co-60 eq = becquerel Cobalt-60, m^2^a = square meter years, N= nitrogen, NO_x_= nitrous oxides, P=phosphate, PM2.5 = particulate matter <2.5 micrometres, SO_2_= sulphur dioxide

**Supplementary Figure 3. Network diagram for laparoscopic scissors, showing freshwater ecotoxicity impact drivers (10% cut-off)**

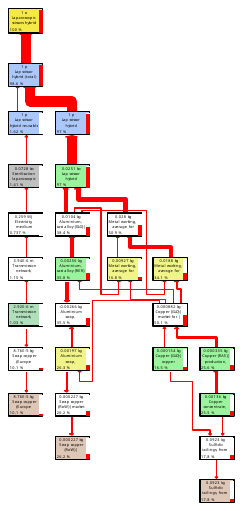

Network diagram for LCA of quantity of hybrid laparoscopic scissors required for a single laparoscopic cholecystectomy (i.e. 1/500 reusable handle, one single-use scissor shaft and blades), showing drivers for the freshwater ecotoxicity category. Each box represents a unit process (only those >10% contribution to the impact are shown), with percentage contribution shown in bottom left, and quantity of the process for the assembly at the top of each box. The arrows represent the flow of materials between processes, and their thickness reflect the relative contribution.

**Supplementary Figure 4. Network diagram for laparoscopic scissors, showing marine ecotoxicity impact drivers (10% cut-off)**

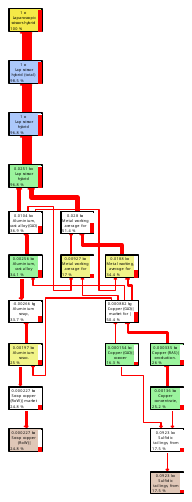

Network diagram for LCA of quantity of hybrid laparoscopic scissors required for a single laparoscopic cholecystectomy (i.e. 1/500 reusable handle, one single-use scissor shaft and blades), showing drivers for the marine ecotoxicity category. Each box represents a unit process (only those >10% contribution to the impact are shown), with percentage contribution shown in bottom left, and quantity of the process for the assembly at the top of each box. The arrows represent the flow of materials between processes, and their thickness reflect the relative contribution.

**Supplementary Figure 5. Network diagram for laparoscopic clip applier, showing marine eutrophication impact drivers (10% cut-off)**

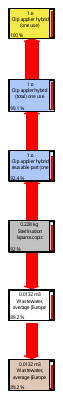

Network diagram for LCA of quantity of hybrid laparoscopic clip applier required for a single laparoscopic cholecystectomy i.e. (1/500 reusable laparoscopic clip applier, one single-use cartridge, showing drivers for the marine eutrophication category. Each box represents a unit process (only those >10% contribution to the impact are shown), with percentage contribution shown in bottom left, and quantity of the process for the assembly at the top of each box. The arrows represent the flow of materials between processes, and their thickness reflect the relative contribution.

**Supplementary Figure 6. Network diagram for hybrid ports, showing marine eutrophication impact drivers (10% cut-off)**

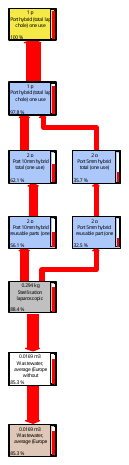

Network diagram for LCA of quantity of hybrid laparoscopic ports required for a single laparoscopic cholecystectomy (i.e. two times 1/500 5 mm and 10 mm cannulas, two times 1/500 5mm and 10mm trocars, one single-use 5 mm duckbill valve, one single-use 5-12 mm universal seal), showing drivers for the marine eutrophication impact category. Each box represents a unit process (only those >10% contribution to the impact are shown), with percentage contribution shown in bottom left, and quantity of the process for the assembly at the top of each box. The arrows represent the flow of materials between processes, and their thickness reflect the relative contribution.

**Supplementary Figure 7. Network diagram for laparoscopic scissors, showing ionising radiation impact drivers (30% cut-off)**

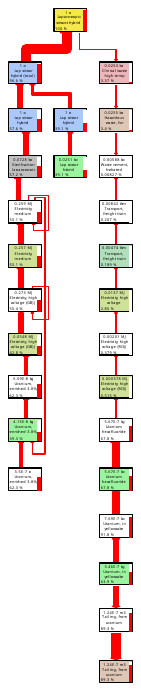

Network diagram for LCA of quantity of hybrid laparoscopic scissors required for a single laparoscopic cholecystectomy (i.e. 1/500 reusable handle, one single-use scissor shaft and blades), showing drivers for the ionising radiation category. Each box represents a unit process (only those >30% contribution to the impact are shown), with percentage contribution shown in bottom left, and quantity of the process for the assembly at the top of each box. The arrows represent the flow of materials between processes, and their thickness reflect the relative contribution.

**Supplementary Figure 8. Network diagram for hybrid ports, showing ionising radiation impact drivers (30% cut-off)**

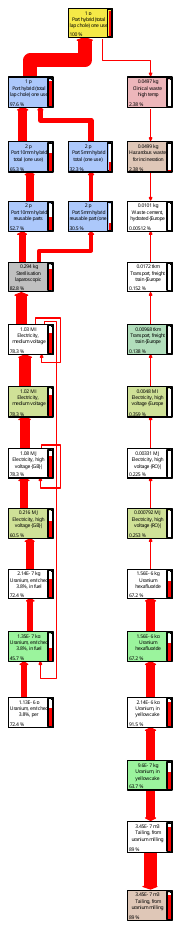

Network diagram for LCA of quantity of hybrid laparoscopic ports required for a single laparoscopic cholecystectomy (i.e. two times 1/500 5 mm and 10 mm cannulas, two times 1/500 5mm and 10mm trocars, one single-use 5 mm duckbill valve, one single-use 5-12 mm universal seal), showing drivers for the marine eutrophication impact category. Each box represents a unit process (only those >10% contribution to the impact are shown), with percentage contribution shown in bottom left, and quantity of the process for the assembly at the top of each box. The arrows represent the flow of materials between processes, and their thickness reflect the relative contribution.

**Supplementary Figure 9. Environmental impact (endpoint categories) of hybrid vs single-use laparoscopic clip applier, scissors, and ports**

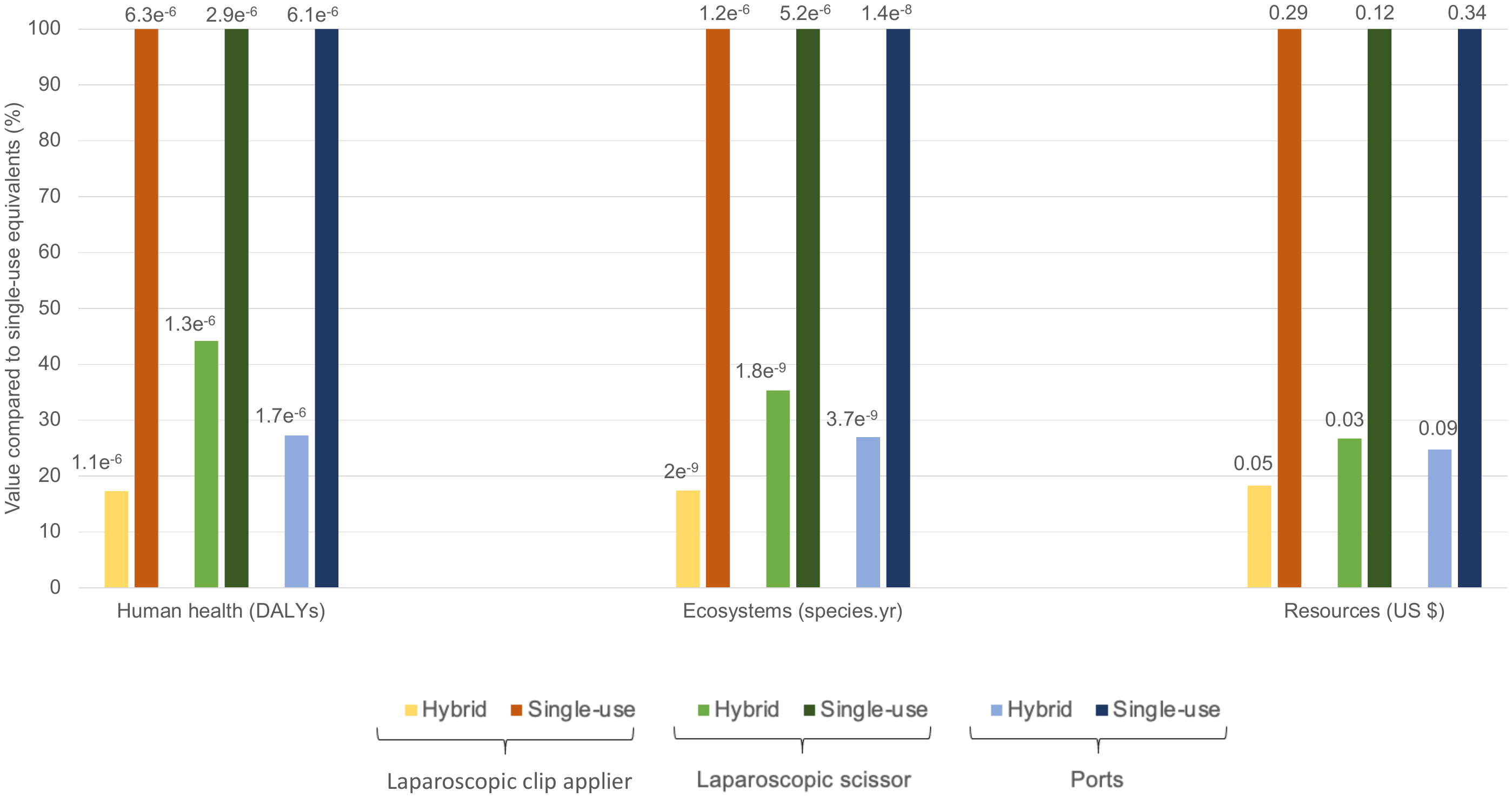

Modelled per use of one laparoscopic clip applier, one laparoscopic scissor, two 5mm ports, and two 10-11mm ports, comparing hybrid with single-use equivalents. Environmental impact compared as proportion (%) of single-use equivalents. Number above bars relate to endpoint category absolute figures for product, measured in disability adjusted life years (DALYs), loss of local species per year (species.year), and extra costs involved for future mineral and fossil resource extraction (US $).

**Supplementary Figure 10. Impact of number of uses of reusable component of hybrid laparoscopic clip applier, scissors, and ports on carbon footprint**

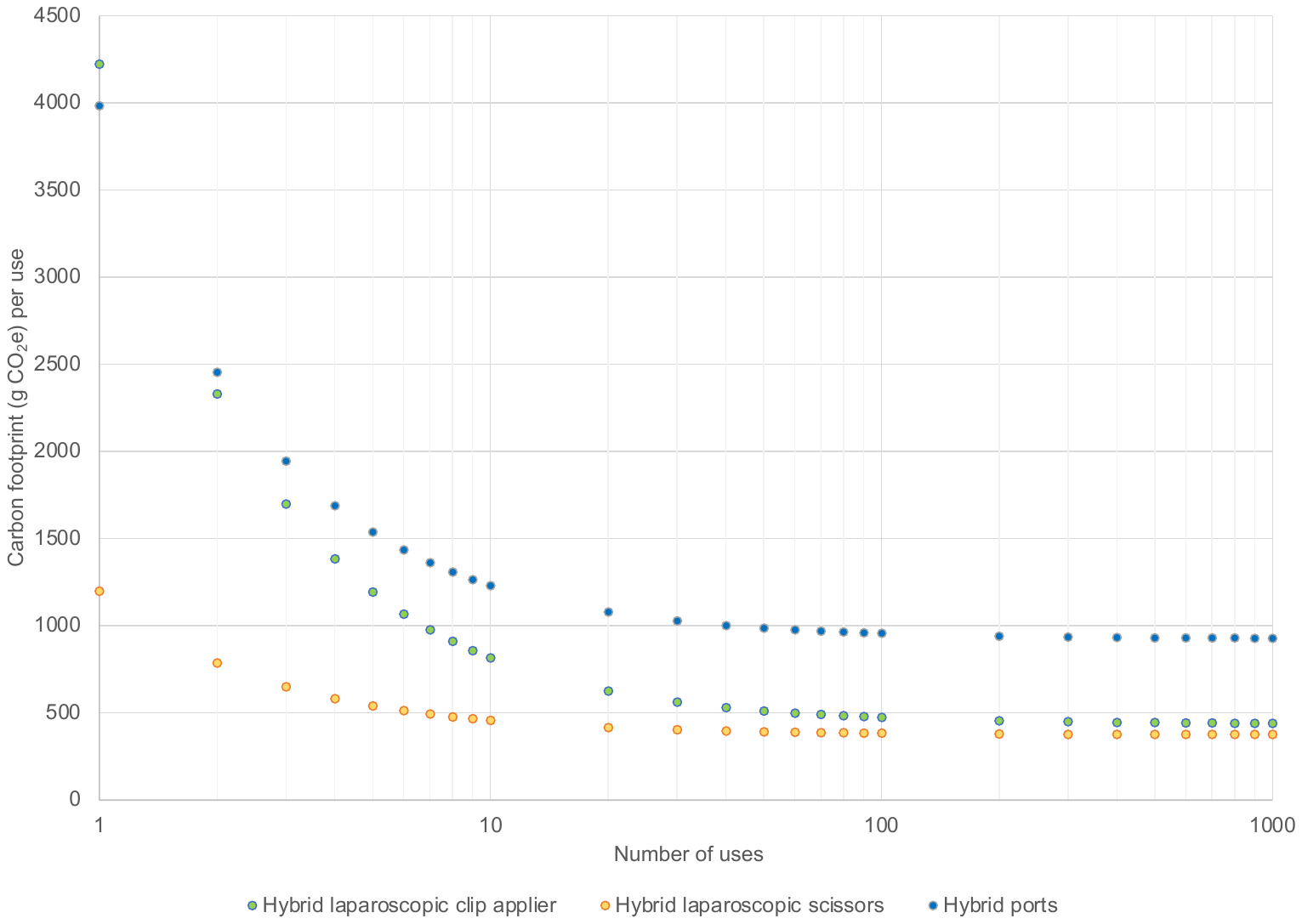

Modelled per use of one hybrid laparoscopic clip applier, one hybrid laparoscopic scissor, two 5mm hybrid ports, and two 10-11mm hybrid ports
