## Supplementary table for "Environmental impact and life cycle financial cost of hybrid (reusable/ single-use) instruments versus single-use equivalents in laparoscopic cholecystectomy"

**Supplementary Table 1: transport assumptions.** Assumes 80km travel by road within site of manufacture country and UK (via heavy goods vehicle, with first and last 8km via courier). Alternative scenarios assuming shipping direct to London Gateway port. * Air freight from California/ Massachusetts (USA) to Amersfoort (Netherlands), to UK via road.

| **Product** | **Site of manufacture** | **Overseas travel** | **Overseas distance (km)** | **UK travel** | **UK travel distance (km)** |
| --- | --- | --- | --- | --- | --- |
|  |  | **(*alternative scenario)*** | **(*alternative scenario)*** |  |  |
| Hybrid laparoscopic clip applier | Massachusetts (USA) | Heavy goods vehicle  Courier  Air freight* | 492  16  5,633 | Heavy goods vehicle  Courier | 64  16 |
| Single-use laparoscopic clip applier | Ohio (USA) | Heavy goods vehicle  Courier  Air freight | 64  16  6,437 | Heavy goods vehicle  Courier | 64  16 |
|  |  | *Heavy goods vehicle*  *Courier*  *Shipping (from Philadelphia)* | *781*  *16*  *5650* |  |  |
| Hybrid laparoscopic scissor | Leeds (UK) | N/A | N/A | Heavy goods vehicle  Courier | 64  16 |
| Single-use laparoscopic scissor | California (USA) | Heavy goods vehicle  Courier  Air freight* | 492  16  8,851 | Heavy goods vehicle  Courier | 64  16 |
|  |  | *Heavy goods vehicle*  *Courier*  *Shipping* | *64*  *16*  *15039* |  |  |
| Hybrid ports | Leeds (UK) | N/A | N/A | Heavy goods vehicle  Courier | 64  16 |
| Single-use ports | California (USA) | Heavy goods vehicle  Courier  Air freight* | 492  16  8,851 | Heavy goods vehicle  Courier | 64  16 |
|  |  | *Heavy goods vehicle*  *Courier*  *Shipping* | *64*  *16*  *15039* |  |  |

Supplementary Table 2: Parameters for decontamination of reusable laparoscopic equipment. Data based on research by the author group previously presented at the International Life Cycle Innovation Conference.(1) kWh= kilowatt hours, m^3^= meters cubed, kg= kilograms

a) Parameters for decontamination of reusable laparoscopic general set, used to house reusable components of hybrid instruments.

| Process | Sub-process | Input | Amount (per laparoscopic set decontaminated) | | |
| --- | --- | --- | --- | --- | --- |
|  |  |  | Weight (g) | Assumed number of uses | Total per use |
| Sterile barrier system | Basket | Stainless steel | 939.12 | 116 | 8.09 g |
|  | Container | Aluminium | 2629.88 | 1000 | 2.63 g |
|  | Container lid | Polyphenylsulfone | 1659.99 | 1000 | 1.66 g |
|  | Identification tag | High Density Polyethylene | 8.21 | 46 | 0.18 g |
|  | Filter paper | Paper | 3.55 | 1 | 8.4 g |
|  | Kit list |  | 4.85 | 1 |  |
|  | Laparoscopic instrument rack | Stainless steel | 937.84 | 500 | 1.88 g |
|  |  | Rubber | 165.50 | 500 | 0.33 g |
|  | Tamper proof tags | Polypropylene granulate | 1.78 | 1 | 1.78 g |
| Decontamination (washing/ disinfection and sterilisation of instrument set) | Detergent | Sodium hydroxide in 50% solution state |  | | 1.85 g |
|  |  | Potassium hydroxide |  |  | 0.93 g |
|  | Electricity | Electricity (UK) |  |  | 1.26 kWh |
|  | Fuel | Natural gas (UK) |  |  | 0.3 m^3^  =11.51 MJ  =3.2 kWh |
|  | Water | Tap water (UK) |  |  | 76.2 kg |
|  |  | Wastewater (UK) |  |  |  |

b) Parameters for decontamination of laparoscopic clip applier as individually wrapped item in single-use flexible pouch

| Process | Sub-process | Input | Amount (per laparoscopic clip applier decontaminated) |
| --- | --- | --- | --- |
| Sterile barrier system | Flexible pouch  (single-use double wrap) | Paper | 20.92 g |
|  |  | Polyethylene | 30.84 g |
|  | Identification tag  (reused as above) | High Density Polyethylene | 0.18 g |
| Decontamination (washing and disinfection) | Detergent | Sodium hydroxide in 50% solution state | 0.46 g |
|  |  | Potassium hydroxide | 0.23 g |
|  | Electricity | Electricity (UK) | 0.32 kWh |
|  | Fuel | Natural gas (UK) | 0.075 m^3^  =2.88 MJ  =0.80 kWh |
|  | Water | Tap water (UK) | 19 kg |
|  |  | Wastewater (UK) |  |

1. Rizan C, Bhutta M, Reed M, Lillywhite R (2020). The carbon footprint of processing reusable surgical instruments. Life Cycle Innovation Conference; Berlin, Germany. Available at: <https://fslci.org/groups/lcic-2020/forum/topic/discussion-session-13-sustainable-pathways-for-decarbonization/>. Accessed 27 Jan 2021.

**Supplementary Table 3: Life cycle inventory processes chosen.** Ecoinvent (version 3.6)- allocation, cut-off by classification- unit library was selected for all processes, aside from where unavailable, Industry data 2.2 library was used.

| **Material** | **Process Name** | **Used for process/ item; comments** |
| --- | --- | --- |
| Aluminium | Aluminium alloy, metal matrix composite {GLO}\| market for \| Cut-off, U | Sterile barrier system for reusable instrument decontamination |
|  | Aluminium, cast alloy {GLO}\| market for \| Cut-off, U | Laparoscopic scissors-hybrid (single-use component) |
|  | Aluminium, wrought alloy {GLO}\| market for \| Cut-off, U | 5 mm port- hybrid (single-use component), 10 mm port- hybrid (single-use component); used for foil |
| Brass | Brass {RoW}\| market for brass \| Cut-off, U | 5 mm port- hybrid (reusable component) |
| Chromium | Chromium {GLO}\| market for \| Cut-off, U | 5 mm port- hybrid (reusable component) |
| Copper | Copper {GLO}\| market for \| Cut-off, U | Laparoscopic scissors-hybrid (reusable component), laparoscopic scissors- single-use |
| Cardboard (corrugated) | Corrugated board box {RoW}\| market for corrugated board box \| Cut-off, U | Clip applier-hybrid (reusable component) |
| Cardboard (boxboard) | Folding boxboard/chipboard {GLO}\| market for \| Cut-off, U | Clip applier-single-use, laparoscopic scissors-hybrid (reusable component) |
| High density polyethylene | Polyethylene, high density, granulate {GLO}\| market for \| Cut-off, U | Clip applier-hybrid (single-use component), clip applier-single-use, laparoscopic scissors-hybrid (single-use component), laparoscopic scissors- single-use, 5 mm port- hybrid (reusable and single-use component), 5 mm port- single-use, 10 mm port- hybrid (single-use component), 10 mm port- hybrid (reusable components), 11 mm port- single-use, sterile barrier system for reusable instrument decontamination |
| Liquid resins | Liquid epoxy resins E | Clip applier-hybrid (reusable component); used for liquid-crystal polymer |
| Low density polyethylene | Polyethylene, low density, granulate {GLO}\| market for \| Cut-off, U | Clip applier-hybrid (single-use component), 5 mm port- single-use, 11 mm port- single-use; used for polyolefin |
|  | Packaging film, low density polyethylene {GLO}\| market for \| Cut-off, U | Clip applier decontamination in flexible pouch |
| Nickel | Nickel, 99.5% {GLO}\| market for \| Cut-off, U | Laparoscopic scissors-hybrid (reusable component), laparoscopic scissors- single-use, 5 mm port- hybrid (reusable component) |
| Nylon | Nylon 6-6 {RoW}\| market for nylon 6-6 \| Cut-off, U | Clip applier-single-use, laparoscopic scissors- single-use, 5 mm port- hybrid (single-use component), 5 mm port- single-use, 10 mm port- hybrid (single-use component), 11 mm port- single-use |
| Paper | Kraft paper, bleached {GLO}\| market for \| Cut-off, U | Clip applier-hybrid (reusable and single-use component), clip applier- single-use, laparoscopic scissor-hybrid (reusable component), laparoscopic scissor-single-use, 5 mm port- hybrid (reusable components), 10 mm port- hybrid (reusable components), sterile barrier system for reusable instrument decontamination |
| Polycarbonate | Polycarbonate {GLO}\| market for \| Cut-off, U | Clip applier-hybrid (single-use component), clip applier-single-use, laparoscopic scissors- single-use, 5 mm port- single-use, 10 mm port- hybrid (single-use component), 11 mm port- single-use |
| Polyester | Polyester resin, unsaturated {RoW}\| market for polyester resin, unsaturated \| Cut-off, U | Laparoscopic scissors- single-use, 10 mm port- hybrid (single-use component) |
| Polyethylene terephthalate | Polyethylene terephthalate, granulate, amorphous {GLO}\| market for \| Cut-off, U | Clip applier-single-use, 5 mm port- hybrid (single-use component), 10 mm port- hybrid (single-use component) |
| Polyoxymethylene | Polyoxymethylene (POM)/EU-27 | 5 mm port- hybrid (reusable component), 10 mm port- hybrid (reusable component) |
| Polyphenylene sulphide | Polyphenylene sulfide {GLO}\| market for \| Cut-off, U | Clip applier-hybrid (reusable component), laparoscopic scissors-hybrid (reusable and single-use component), 5 mm port- hybrid (reusable component), 10 mm port- hybrid (reusable component); used as substitute for PEEK (polyether ether ketone); laparoscopic general set container lid |
| Polypropylene | Polypropylene, granulate {GLO}\| market for \| Cut-off, U | Clip applier-single-use, laparoscopic scissors- single-use, 5 mm port- single-use, 11 mm port- single-use, sterile barrier system for reusable instrument decontamination |
| Polyurethane | Polyurethane, flexible foam {RoW}\| market for polyurethane, flexible foam \| Cut-off, U | Clip applier hybrid-reusable component, 11 mm port- single-use |
| Polyvinylchloride | Polyvinylchloride, suspension polymerised {GLO}\| market for \| Cut-off, U | Clip applier-single-use, 5 mm port- hybrid (reusable component),10 mm port- hybrid (reusable components) |
| Potassium hydroxide | Potassium hydroxide {GLO}\| market for \| Cut-off, U | Detergent for reusable instrument decontamination |
| Rubber | Synthetic rubber {GLO}\| market for \| Cut-off, U | Laparoscopic instrument rack for reusable instrument decontamination |
| Silicone | Silicone product {RoW}\| market for silicone product \| Cut-off, U | Laparoscopic scissors-hybrid (single-use component), laparoscopic scissors- single-use, 5 mm port- hybrid (single-use component), 5 mm port- single-use, 10 mm port- hybrid (single-use component), 11 mm port- single-use |
| Sodium hydroxide | Sodium hydroxide, without water, in 50% solution state {GLO}\| market for \| Cut-off, U | Detergent for reusable instrument decontamination |
| Stainless steel | Steel, chromium steel 18/8 {GLO}\| market for \| Cut-off, U | Clip applier-hybrid (reusable and single-use component), clip applier-single-use, laparoscopic scissors-hybrid (reusable and single-use component), laparoscopic scissors- single-use, 5 mm port- hybrid (reusable component), 10 mm port- hybrid (reusable component), 10 mm port- hybrid (reusable component), 11 mm port- single-use, sterile barrier system for reusable instrument decontamination, laparoscopic instrument rack for reusable instrument decontamination |
| Titanium | Titanium, primary {GLO}\| market for \| Cut-off, U | Clip applier-hybrid (single-use component), clip applier-single-use |
| Zinc | Zinc {GLO}\| market for \| Cut-off, U | Laparoscopic scissors-hybrid (reusable component), laparoscopic scissors- single-use |
| **Energy** | **Process Name** | **Used for process/ item; comments** |
| Electricity (UK) | Electricity, medium voltage {GB}\| market for \| Cut-off, U | Reusable instrument decontamination |
| **Processing** | **Process name** | **Used for process/ item; comments** |
| Plastic product manufacturing | Injection moulding {GLO}\| market for \| Cut-off, U | Manufacture of all plastics; auxiliaries and energy demand for conversion of plastics via injection moulding |
| Metal product manufacturing | Metal working, average for metal product manufacturing {GLO}\| market for \| Cut-off, U | Manufacture of all metals; manufacturing processes to make a semi-manufactured product into a final product, including average values for the processing of metals by machines and factory infrastructure and operation, plus steel input for loss during processing |
| Production of steam | Process steam from natural gas, heat plant, consumption mix, at plant, MJ GB S | Reusable instrument decontamination |
| **Transportation** | **Process Name** | **Used for process/ item; comments** |
| Air freight | Transport, freight, aircraft, long haul {GLO}\| market for transport, freight, aircraft, long haul \| Cut-off, U | Transport via air freight |
| Courier | Transport, freight, light commercial vehicle {RER}\| market group for transport, freight, light commercial vehicle \| Cut-off, U | Transport via courier |
| Heavy goods vehicle | Transport, freight, lorry, unspecified {RER}\| market for transport, freight, lorry, unspecified \| Cut-off, U | Transport via heavy goods vehicle |
| Shipping | Transport, freight, sea, container ship {GLO}\| market for transport, freight, sea, container ship \| Cut-off, U | Alternative transport scenario- shipping |
| **Water** | **Process Name** | **Used for process/ item; comments** |
| Tap water | Tap water {RER}\| market group for \| Cut-off, U | Reusable instrument decontamination |
| Wastewater | Wastewater, average {Europe without Switzerland}\| market for wastewater, average \| Cut-off, U | Reusable instrument decontamination |
| **Waste** | **Process Name** | **Used for process/ item; comments** |
| Hazardous waste | Hazardous waste, for incineration {Europe without Switzerland}\| treatment of hazardous waste, hazardous waste incineration \| Cut-off, U | All waste streams |

**Supplementary Table 4. Contributions of processes to carbon footprint of hybrid versus single-use laparoscopic clip applier, scissors, and ports.** Contributions of processes to carbon footprint of one laparoscopic clip applier, one laparoscopic scissor, two 5mm ports, and two 10-11mm ports (one use; number required to perform a single laparoscopic cholecystectomy

| **Component** | **Process** | **Carbon footprint (g CO_2_)** | | | | | |
| --- | --- | --- | --- | --- | --- | --- | --- |
|  |  | **Laparoscopic clip applier** | | **Laparoscopic scissors** | | **Ports** | |
|  |  | Hybrid | Single-use | Hybrid | Single-use | Hybrid | Single-use |
| Reusable component | Raw material extraction and manufacture | 4.37 | N/A | 1.27 | N/A | 4.58 | N/A |
|  | Transportation | 2.05 |  | 0.01 |  | 0.04 |  |
|  | Decontamination | 247 |  | 79 |  | 319 |  |
|  | Waste | 1.15 |  | 0.37 |  | 1.49 |  |
| Single-use component | Raw material extraction and manufacture | 112 | 1,342 | 232 | 660 | 481 | 2,122 |
|  | Transportation | 42 | 923 | 2 | 324 | 2 | 823 |
|  | Waste | 36 | 294 | 64 | 154 | 125 | 550 |
| Total | | **445** | **2,559** | **378** | **1,139** | **933** | **3,495** |

**Supplementary Table 5: Alternative decontamination assumptions; environmental impact (midpoint categories) per use of hybrid laparoscopic clip applier, scissors, and ports.** Here, we assumed that the laparoscopic clip applier was individually prepared and housed in a flexible polyethylene pouch (double wrapped), and that the total weight of items contained within the laparoscopic general set was reduced to 1.1kg, and the decontamination of the surgical scissors and ports were apportioned accordingly. Environmental impacts (midpoint categories) measured using life cycle assessment and modelled on one laparoscopic clip applier, one laparoscopic scissor, two 5mm ports, and two 10-11mm ports (one use; number required to perform a single laparoscopic cholecystectomy), comparing hybrid with single-use equivalents. 1,4-DCB =dichlorobenzene, CFC11= Trichlorofluoromethane, CO_2_e= carbon dioxide equivalents, Cu= copper, eq= equivalents, Bq Co-60 eq = becquerel Cobalt-60, m^2^a = square meter years, N= nitrogen, NO_x_= nitrous oxides, P=phosphate, PM2.5 = particulate matter <2.5 micrometres, SO_2_= sulphur dioxide

| **Impact category** | **Unit** | **Hybrid laparoscopic clip applier** | **Hybrid laparoscopic scissors** | **Hybrid ports** |
| --- | --- | --- | --- | --- |
| Global warming | g CO2e | 1650 | 394 | 999 |
| Stratospheric ozone depletion | g CFC11 eq | 0.0006 | 0.0001 | 0.0004 |
| Ionizing radiation | Bq Co-60 eq | 256 | 32 | 92 |
| Ozone formation, Human health | g NOx eq | 2.90 | 0.81 | 1.50 |
| Fine particulate matter formation | g PM2.5 eq | 1.71 | 0.79 | 0.93 |
| Ozone formation, Terrestrial ecosystems | g NOx eq | 3.00 | 0.83 | 1.55 |
| Terrestrial acidification | g SO2 eq | 3.50 | 1.47 | 2.18 |
| Freshwater eutrophication | g P eq | 0.37 | 0.17 | 0.18 |
| Marine eutrophication | g N eq | 0.38 | 0.04 | 0.14 |
| Terrestrial ecotoxicity | g 1,4-DCB | 5125 | 5656 | 1283 |
| Freshwater ecotoxicity | g 1,4-DCB | 59 | 97 | 19 |
| Marine ecotoxicity | g 1,4-DCB | 78 | 122 | 25 |
| Human carcinogenic toxicity | g 1,4-DCB | 72 | 66 | 45 |
| Human non-carcinogenic toxicity | g 1,4-DCB | 1188 | 960 | 422 |
| Land use | m2a crop eq | 0.13 | 0.01 | 0.03 |
| Mineral resource scarcity | g Cu eq | 11 | 14 | 3 |
| Fossil resource scarcity | g oil eq | 628 | 106 | 284 |
| Water consumption | m3 | 0.0172 | 0.0029 | 0.0086 |

**Supplementary Table 6: Alternative energy supply assumptions; environmental impact (midpoint categories) per use of hybrid laparoscopic clip applier, scissors, and ports.** Here, we assumed that Australian electricity was used for decontamination of reusable components. Environmental impacts (midpoint categories) measured using life cycle assessment and modelled on one laparoscopic clip applier, one laparoscopic scissor, two 5mm ports, and two 10-11mm ports (one use; number required to perform a single laparoscopic cholecystectomy), comparing hybrid with single-use equivalents. 1,4-DCB =dichlorobenzene, CFC11= Trichlorofluoromethane, CO_2_e= carbon dioxide equivalents, Cu= copper, eq= equivalents, Bq Co-60 eq = becquerel Cobalt-60, m^2^a = square meter years, N= nitrogen, NO_x_= nitrous oxides, P=phosphate, PM2.5 = particulate matter <2.5 micrometres, SO_2_= sulphur dioxide

| **Impact category** | **Unit** | **Hybrid laparoscopic clip applier** | **Hybrid laparoscopic scissors** | **Hybrid ports** |
| --- | --- | --- | --- | --- |
| Global warming | g CO2e | 579 | 421 | 1,105 |
| Stratospheric ozone depletion | g CFC11 eq | 0.0003 | 0.0002 | 0.0006 |
| Ionizing radiation | Bq Co-60 eq | 11 | 13 | 19 |
| Ozone formation, Human health | g NOx eq | 1.16 | 0.87 | 1.76 |
| Fine particulate matter formation | g PM2.5 eq | 0.78 | 0.83 | 1.09 |
| Ozone formation, Terrestrial ecosystems | g NOx eq | 1.18 | 0.89 | 1.80 |
| Terrestrial acidification | g SO2 eq | 1.66 | 1.59 | 2.69 |
| Freshwater eutrophication | g P eq | 0.47 | 0.28 | 0.61 |
| Marine eutrophication | g N eq | 0.11 | 0.04 | 0.14 |
| Terrestrial ecotoxicity | g 1,4-DCB | 4,010 | 5,639 | 1,215 |
| Freshwater ecotoxicity | g 1,4-DCB | 45 | 100 | 28 |
| Marine ecotoxicity | g 1,4-DCB | 59 | 126 | 38 |
| Human carcinogenic toxicity | g 1,4-DCB | 62 | 71 | 65 |
| Human non-carcinogenic toxicity | g 1,4-DCB | 918 | 1,062 | 831 |
| Land use | m2a crop eq | 0.01 | 0.01 | 0.01 |
| Mineral resource scarcity | g Cu eq | 9 | 14 | 3 |
| Fossil resource scarcity | g oil eq | 162 | 108 | 294 |
| Water consumption | m3 | 0.003 | 0.003 | 0.008 |

**Supplementary Table 7: Alternative overseas transportation assumptions; environmental impact (midpoint categories) per use of single-use laparoscopic clip applier, scissors, and ports.** Here, we assumed that shipping was used for overseas transportation. Environmental impacts (midpoint categories) measured using life cycle assessment and modelled on one laparoscopic clip applier, one laparoscopic scissor, two 5mm ports, and two 10-11mm ports (one use; number required to perform a single laparoscopic cholecystectomy), comparing hybrid with single-use equivalents. 1,4-DCB =dichlorobenzene, CFC11= Trichlorofluoromethane, CO_2_e= carbon dioxide equivalents, Cu= copper, eq= equivalents, Bq Co-60 eq = becquerel Cobalt-60, m^2^a = square meter years, N= nitrogen, NO_x_= nitrous oxides, P=phosphate, PM2.5 = particulate matter <2.5 micrometres, SO_2_= sulphur dioxide

| **Impact category** | **Unit** | **Single-use laparoscopic clip applier** | **Single-use laparoscopic scissors** | **Single-use ports** |
| --- | --- | --- | --- | --- |
| Global warming | g CO2e | 1,727 | 837 | 2,728 |
| Stratospheric ozone depletion | g CFC11 eq | 0.0006 | 0.0004 | 0.0011 |
| Ionizing radiation | Bq Co-60 eq | 72 | 22 | 52 |
| Ozone formation, Human health | g NOx eq | 4.28 | 1.88 | 4.91 |
| Fine particulate matter formation | g PM2.5 eq | 3.39 | 1.69 | 2.96 |
| Ozone formation, Terrestrial ecosystems | g NOx eq | 4.39 | 1.93 | 5.07 |
| Terrestrial acidification | g SO2 eq | 6.41 | 3.81 | 7.24 |
| Freshwater eutrophication | g P eq | 0.61 | 0.25 | 0.42 |
| Marine eutrophication | g N eq | 0.06 | 0.05 | 0.07 |
| Terrestrial ecotoxicity | g 1,4-DCB | 19,061 | 8,342 | 2,622 |
| Freshwater ecotoxicity | g 1,4-DCB | 174 | 90 | 37 |
| Marine ecotoxicity | g 1,4-DCB | 227 | 116 | 49 |
| Human carcinogenic toxicity | g 1,4-DCB | 201 | 90 | 115 |
| Human non-carcinogenic toxicity | g 1,4-DCB | 2,717 | 1,319 | 843 |
| Land use | m2a crop eq | 0.16 | 0.02 | 0.02 |
| Mineral resource scarcity | g Cu eq | 38.78 | 18.96 | 3.46 |
| Fossil resource scarcity | g oil eq | 514 | 216 | 689 |
| Water consumption | m3 | 0.014 | 0.008 | 0.020 |

**Supplementary Table 8: Alternative port set up assumptions; environmental impact (midpoint categories) per use of hybrid vs single-use ports.** Hybrid model based upon three 5mm ports, one 10mm port; single-use model based upon three 5mm ports, and one 11mm port. 1,4-DCB =dichlorobenzene, CFC11= Trichlorofluoromethane, CO_2_e= carbon dioxide equivalents, Cu= copper, eq= equivalents, Bq Co-60 eq = becquerel Cobalt-60, m^2^a = square meter years, N= nitrogen, NO_x_= nitrous oxides, P=phosphate, PM2.5 = particulate matter <2.5 micrometres, SO_2_= sulphur dioxide

| **Impact category** | **Unit** | **Hybrid ports** | **Single-use ports** |
| --- | --- | --- | --- |
| Global warming | g CO2e | 635 | 3,613 |
| Stratospheric ozone depletion | g CFC11 eq | 0.0003 | 0.0013 |
| Ionizing radiation | Bq Co-60 eq | 65 | 60 |
| Ozone formation, Human health | g NOx eq | 0.97 | 8.41 |
| Fine particulate matter formation | g PM2.5 eq | 0.59 | 3.58 |
| Ozone formation, Terrestrial ecosystems | g NOx eq | 0.99 | 8.60 |
| Terrestrial acidification | g SO2 eq | 1.39 | 9.13 |
| Freshwater eutrophication | g P eq | 0.12 | 0.45 |
| Marine eutrophication | g N eq | 0.10 | 0.08 |
| Terrestrial ecotoxicity | g 1,4-DCB | 900 | 3,997 |
| Freshwater ecotoxicity | g 1,4-DCB | 13 | 39 |
| Marine ecotoxicity | g 1,4-DCB | 17 | 53 |
| Human carcinogenic toxicity | g 1,4-DCB | 29 | 125 |
| Human non-carcinogenic toxicity | g 1,4-DCB | 289 | 1,032 |
| Land use | m2a crop eq | 0.02 | 0.03 |
| Mineral resource scarcity | g Cu eq | 2 | 3 |
| Fossil resource scarcity | g oil eq | 186 | 970 |
| Water consumption | m3 | 0.0060 | 0.0220 |
